# Chronic pulmonary aspergillosis incidence in newly detected pulmonary tuberculosis cases during follow-up

**DOI:** 10.1101/2024.02.28.24303416

**Authors:** Dhouli Jha, Umesh Kumar, Ved Prakash Meena, Prayas Sethi, Amandeep Singh, Neeraj Nischal, Pankaj Jorwal, Surabhi Vyas, Gagandeep Singh, Immaculata Xess, Urvashi B Singh, Sanjeev Sinha, Anant Mohan, Naveet Wig, Sushil Kumar Kabra, Animesh Ray

**Author notes:** Corresponding author: Dr Animesh Ray Additional Professor Department of Medicine, All India Institute of Medical Sciences New Delhi-110029, India, / 26588641, Email ID.

## Abstract

**Background:** Chronic pulmonary aspergillosis (CPA) is known to complicate patients with post-tubercular lung disease. However, some evidence suggests that CPA might co-exist in patients with newly-diagnosed pulmonary tuberculosis (P.TB) at diagnosis and also develop during therapy. The objective of this study was to confirm the presence of CPA in newly diagnosed P.TB at baseline and at end-of-therapy.

**Materials & Methods:** This prospective longitudinal study included newly diagnosed P.TB patients, followed up at third month and end-of-therapy with symptom assessment, anti-*Aspergillus* IgG antibody and imaging of chest for diagnosing CPA.

**Results:** We recruited 255 patients at baseline out of which 158 (62%) completed their follow-up. Anti-*Aspergillus* IgG was positive in 11.1% at baseline and 27.8% at end-of-therapy. Overall, proven CPA was diagnosed in 7% at baseline and 14.5% at end-of-therapy. Around 6% patients had evidence of aspergilloma in CT chest at the end-of-therapy.

**Conclusions:** CPA can be present in newly diagnosed P.TB patients at diagnosis and also develop during anti-tubercular treatment. Patients with persistent symptoms or developing new symptoms during treatment for P.TB should be evaluated for CPA.

## Introduction

Chronic pulmonary aspergillosis (CPA) is a chronic fungal infection of the lung resulting in myriad clinical manifestations,^1^ usually diagnosed by compatible clinical features along with radio-microbiological characteristics. The presence of IgG anti-*Aspergillus* is a cornerstone of diagnosis ^2^ Early diagnosis of this condition is important as despite antifungal treatment mortality can be substantial.^3^

CPA typically occurs due to colonization and subsequent infection by *Aspergillus* in the background of structural lung disease. In tuberculosis (TB)-endemic countries the commonest predisposing lung condition is often post-tubercular lung disease (PTLD) while in other areas, conditions like chronic obstructive pulmonary disease (COPD) may be more frequent.^4, 5^ The relationship between TB and CPA however extends beyond the entity of PTLD. Due to similar clinico-radiological features, CPA may often be misdiagnosed as sputum-negative pulmonary tuberculosis (P.TB).^6^ Further, a recent systematic review had identified the co-existence (possibly including colonization) of *Aspergillus* fungal bodies in patients suffering from P.TB in up to 33.3% of cases– however without considering the presence of antibodies or diagnosis of CPA.^7^

Recently, there has been interest in the possibility of co-occurrence of CPA in newly diagnosed P.TB patients at baseline and also during therapy. An Indonesian study had reported CPA in 6% of newly diagnosed sputum-positive and sputum-negative P.TB patients which increased to 8% by the end-of-therapy.^8^ A retrospective study from Korea reported on the changes in anti-*Aspergillus* IgG antibody from the time of diagnosis of P.TB till after treatment. ^9^ In this study, where CPA was diagnosed in 2.9% of patients at a median of 13.5 months after treatment completion, they noticed significant shift in antibody-titer (from positive to negative or opposite thereof) in 11.6% of patients. This raised the possibility of self-resolving CPA (vis-à-vis chronic progressive pulmonary aspergillosis)^10^. However, the said studies suffered from several methodological inadequacies like retrospective design, CT thorax in selected patients as well as significant loss to follow-up. The present study was designed to estimate the burden of anti-*Aspergillus* IgG positivity and CPA in newly-diagnosed P.TB patients both at baseline and end-of-therapy with the aim of avoiding the limitations mentioned above.

## Methods

### Study population

Between April 2022 to December 2022, consecutive newly diagnosed P.TB patients within seven days of treatment initiation, presenting to a TB clinic in south Delhi were enrolled after obtaining written consent. In this study patients with previous history of TB, underlying chronic lung condition, solid organ transplant recipients, active malignancy or undergoing radiotherapy, pregnant female, HIV-positive patients and patients below 14 years of age were excluded. Ethical permission was taken from Institute Ethics Committee for the conduct of this study (IECPG-311/27.04.2022).

### Assessment and investigations at different time points

Patients were assessed at different time points from the beginning of recruitment (baseline, after third month and after sixth month of therapy initiation i.e., after completion of antitubercular therapy for most patients). For all these time points, patients underwent symptom screening, anti-*Aspergillus* IgG levels and quality of life scores with SGRQ score (St Georges Respiratory Questionnaire)^11^. Lung imaging was done at two time points-at baseline (chest radiograph) and at the end-TB therapy (high resolution computed tomography of thorax/HRCT thorax or chest radiograph). Sputum examination for fungal stain/ fungal culture were done whenever possible at two time points-baseline and end-TB therapy.

The data were first recorded in pre-designed case records form and transferred to digital format by trained personnel. The lung images were assessed with the help of standard reporting form containing list of radiological features routinely encountered in post-TB sequelae patients.^12^ The HRCT thorax scans were reported by a trained radiologist with special interest in thoracic radiology (SV) while the radiographs were assessed by a pulmonologist (with more than eight years’ experience in evaluating CPA patients). Anti-*Aspergillus* IgG was assessed by using ImmunoCAP *Asp* IgG assay (Thermo Fisher, Waltham, MA, USA) and a cut off >27 mg/L was considered as positive. Serum galactomannan (GM) was measured by Platelia *Aspergillus* galactomannan ELISA (Bio-Rad Laboratories) and interpreted according to guidance in the 2019 EORTC/MSGERC guidelines.^13^ A GM index >1 was considered positive for serum. Figure 1 describes the study flow plan.

**Figure 1:**
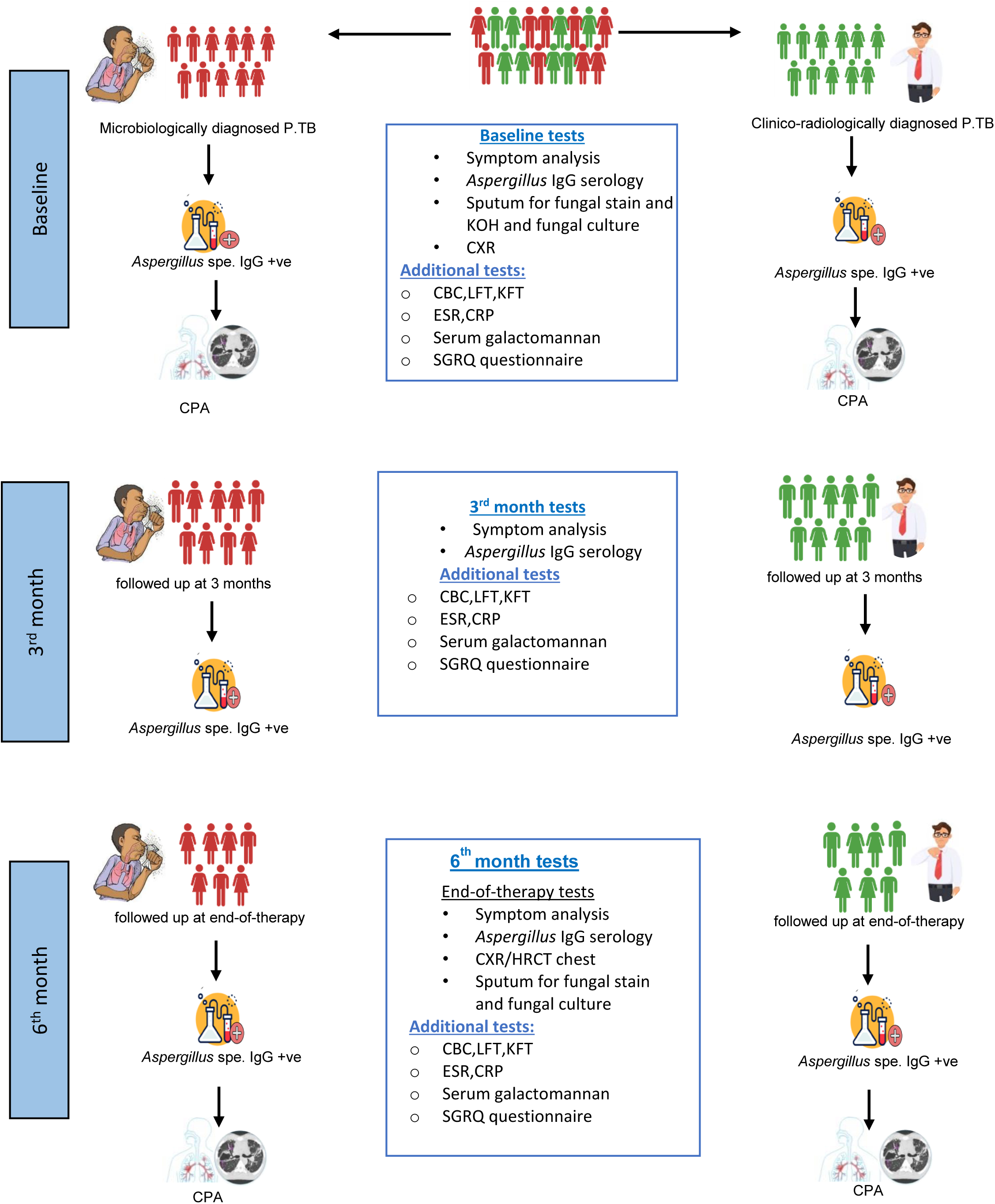
Study flow plan

### Clinical definitions

Chronic pulmonary aspergillosis was diagnosed on the basis of presence of standard criteria^14^.The details of diagnostic criteria are given in supplementary appendix.

At the end-of-therapy, we divided the patients into four groups – ‘never CPA’, ‘persistent CPA’, ‘self-resolving CPA’ ^8^ and ‘new-CPA’ based on their symptoms and investigations (vide supplementary appendix).

### Statistical analysis

Data was analysed using STATA 14.0 version by trained personnel (AR & AD). Qualitative data was presented in frequency and in percentages whereas quantitative data was reported as mean/standard deviation or median (with interquartile range) according to the normality of data distribution. To establish association between qualitative variables, chi square / fisher exact was used while student T test / Wilcoxon signed rank test was used as appropriate for quantitative variables. Value of p< 0.05 was considered as statistically significant.

## Results

Out of the initially recruited 255 patients diagnosed with P.TB, 158 (62%) were followed up until the completion of therapy. At baseline, 160 P.TB patients (63%) were microbiologically confirmed. Sputum ZN staining could be performed in 168 patients (66%) at baseline, with 74% yielding positive results. GeneXpert MTB/RIF tests were done in 165 patients (65%), with 95% positive reports (149 Rifampicin-sensitive, 7 resistant, and 1 indeterminate case). Liquid culture by MGIT was performed in 132 patients (52%) at baseline, with an 86% positivity rate – 90% in microbiologically diagnosed P.TB and 10% in initial clinico-radiologically diagnosed cases.

At baseline, 18 out of 255 patients (7%) met CPA criteria. Clinico-demographic features (Table 1) showed a significantly higher mean age in the CPA group (38.8 vs. 30.9 years, p=0.02). Hemoptysis occurred more in CPA patients (22% in CPA vs. 4% in non-CPA, p=0.011). Baseline SGRQ scores were however similar (58.47 in CPA vs. 57.82 in non-CPA). Radiologically, 26% patients had cavities at baseline (29% in CPA). Pleural thickening was observed in 2 patients (1%), though none were diagnosed as CPA. Among 228 patients with processed sputum, 7 (3%) had *Aspergillus*-positive cultures. *Aspergillus flavus* (n=3, 1.2%) was the commonest of all followed by *Aspergillus fumigatus* (n=2, 0.8%) and *A. niger* (n=2, 0.8%). None with positive cultures met CPA criteria.

**Table 1:** Baseline characteristics of recruited patients.

| Baseline characteristics | All (n=255) | CPA (n=18) | Non-CPA (n=237) | P value |
| --- | --- | --- | --- | --- |
| **Gender** |  |  |  |  |
| Male | 153(60%) | 9(50%) | 144(61%) | 0.369 |
| Female | 102(40%) | 9(50%) | 93(39%) |  |
| **Age (Mean+/-SD)** | 31.5(13.9) | 38.8(19.4) | 30.9(13.2) | **0.02** |
| **Symptomatology (>3months)** |  |  |  |  |
| 1.Fever | 37(15%) | 3(17%) | 34(14%) | 0.507 |
| 2.Cough | 58(23%) | 7(39%) | 51(22%) | 0.217 |
| 3.Fatigue | 47(18%) | 6(33%) | 41(17%) | 0.250 |
| 4.Chest pain | 27(11%) | 3(17%) | 24(10%) | 0.222 |
| 5.Dyspnea | 31(12%) | 5(28%) | 26(11%) | 0.066 |
| 6.Hemoptysis | 13(5%) | 4(22%) | 9(4%) | **0.011** |
| 7.Loss of weight | 41(16%) | 5(28%) | 36(15%) | 0.305 |
| 8.Loss of appetite | 48(19%) | 4(22%) | 44(19%) | 0.340 |
| **Comorbidities** |  |  |  |  |
| Diabetes mellitus | 30(12%) | 3(17%) | 27(11%) | 0.503 |
| Hypertension | 10(4%) | 0 | 10(4%) | 0.386 |
| CKD | 1(0.4%) | 1(6%) | 0 | 0.067 |
| CLD | 1(0.4%) | 0 | 1(0.4%) | 0.788 |
| Smoking | 102(40%) | 5(28%) | 97(41%) | 0.272 |
| *Aspergillus* specific IgG titre | 13.28 (11.6-14.9) | 41.44 (33.7-49.1) | 11.12 (9.8-12.5) | **<0.001** |
| Serum galactomannan (Median with IQR) | 0.329 (0.23-0.475) | 0.209 (0.117-0.326) | 0.338 (0.24-0.496) | 0.898 |
| **Radiology** |  |  |  |  |
| 1.Cavitation | 63(26%) | 5(29%) | 58(26%) | 0.775 |
| 2.Nodules | 192(81%) | 14(82%) | 178(81%) | 1.000 |
| 3.Aspergilloma | 0 | 0 | 0 | .... |
| 4.Pleural thickening | 2(1%) | 0 | 2(1%) | 1.000 |
| 6.Consolidation | 183(76%) | 14(82%) | 169(76%) | 0.769 |
| SGRQ score baseline |  |  |  |  |
| Symptoms | 43.9(21.3) | 42.5(19.9) | 44.1(21.5) | 0.766 |
| Impact | 57.39(18.3) | 55.86(19) | 57.51(18.2) | 0.7125 |
| Activity | 58.69(22.2) | 62.71(22.9) | 58.38(22.2) | 0.426 |
| Total | 57.86 (17.5) | 58.47 (19.3) | 57.82 (17.4) | 0.879 |
| Anemia | 174(67%) | 11(61%) | 163(69%) | 0.484 |
| Elevated CRP | 223(89%) | 16(89%) | 207(89%) | 1.000 |

Five (2%) enrolled patients died before treatment completion. Only one had microbiological evidence of P.TB. Two patients had symptom/s >3 months at baseline. None had raised anti-Aspergillus IgG antibody. On lung imaging, three patients had consolidation, two had nodules, one patient had extensive miliary mottling and one had lung cavity. No deceased patients were categorized as CPA at baseline.

There were eight (3%) drug resistant P.TB patients, two of them were lost to follow-up by the end of study. One had CPA at baseline, which ‘self-resolved’ by the end of six months of anti-tubercular therapy. Two developed ‘new CPA’ at the sixth month follow-up.

At the end-of-TB therapy, 23 (15%) out of 158 assessed patients were diagnosed with CPA (Figure 2a). CPA group had higher rates of persistent cough (43% vs. 16% in non-CPA, p=0.001) and hemoptysis (9% vs. 1% in non-CPA, p=0.05) (Table 2). The overall prevalence of positive anti-*Aspergillus* IgG was 27.8% (44/158), with all CPA patients testing positive, compared to 16.3% in the non-CPA group. Quality of life (QOL) assessed by SGRQ score decreased overall during TB therapy (56.3 at baseline to 37.2 at end-of-therapy). However, in CPA, SGRQ score remained significantly higher than non-CPA (53.4 vs. 34.4, p=0.0017). Radiologically, CPA group exhibited higher frequency of consolidation (39% vs. 16% in non-CPA, p=0.010) and cavitation (61% vs. 31% in non-CPA, p=0.006) by end-of-therapy. Fungal balls were present in CT images of four patients, with only one diagnosed as CPA.

**Figure 2.**
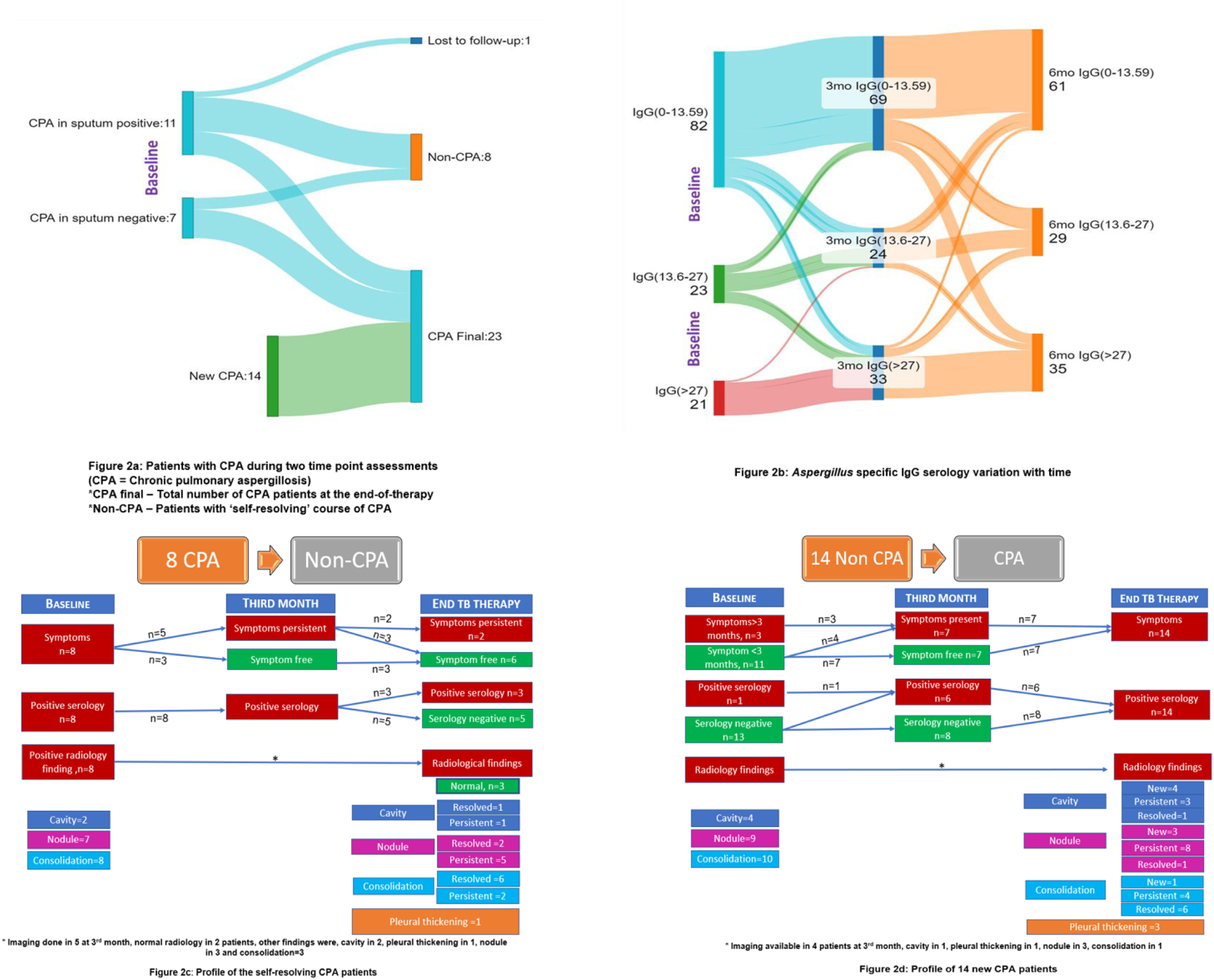
2a: Patients with CPA at two time point assessment 2b: Aspergillus specific IgG serology variation with time 2c and 2d: Clinic0-serological-demographic details for ‘self-resolving’ CPA and ‘new CPA’

**Table 2:** Results for end of TB therapy assessment compared with baseline for all those assessed at both time points, using the CPA categorisation at the second time point.

| End-of-therapy characteristics | Early TB therapy (n=158) | End TB therapy (n=158) | CPA (n=23) | Non-CPA (n=135) | P value* |
| --- | --- | --- | --- | --- | --- |
| Male | 87(55%) | 87(55%) | 11(48%) | 76(56%) | 0.450 |
| Age(Mean+/-SD) | 32.1(14.6) | 32.1(14.6) | 34.4(16.9) | 31.7(14.1) | 0.411 |
| Symptomatology (>3months) |  |  |  |  |  |
| 1.Fever | 27(17%) | 4(3%) | 0(0%) | 4(3%) | 1.000 |
| 2.Cough | 41(26%) | 31(20%) | 10(43%) | 21(16%) | **0.001** |
| 3.Fatigue | 36(23%) | 40(25%) | 8(35%) | 32(24%) | 0.452 |
| 4.Chest pain | 21(13%) | 23(15%) | 6(26%) | 17(13%) | 0.065 |
| 5.Dyspnea | 24(15%) | 52(33%) | 12(52%) | 40(30%) | 0.078 |
| 6.Hemoptysis | 11(7%) | 3(2%) | 2(9%) | 1(1%) | **0.056** |
| 7.Loss of weight | 31(20%) | 9(6%) | 2(9%) | 7(5%) | 0.676 |
| 8.Loss of appetite | 34(22%) | 10(6%) | 2(9%) | 8(6%) | 0.773 |
| *Aspergillus* specific IgG titre | 13.9 (11.6-16.2) | 24.6 (19.7-29.6) | 66.7 (46.9-86.4) | 17.8 (14-21.6) | < 0.001 |
| Serum galactomannan | 0.33 (0.22-0.46) | 0.37 (0.26-0.65) | 0.63 (0.38-0.91) | 0.35 (0.25-0.61) | 0.898 |
| SGRQ score | 56.3 (53.4-59.2) | 37.2 (32.9-41.4) | 53.4 (42.6-64.1) | 34.4 (29.9-38.9) | **0.0017** |
| Radiology |  |  |  |  |  |
| 1.Cavitation | 36(25%) | 53(36%) | 14(61%) | 39(31%) | **0.006** |
| 2.Nodules | 117(80%) | 115(78%) | 20(87%) | 95(76%) | 0.291 |
| 3.Aspergilloma ^ | 0 | 4(3%) | 1(4%) | 3(2%) | 0.495 |
| 4.Pleural thickening | 1(1%) | 24(16%) | 5(22%) | 19(15%) | 0.434 |
| 5.Bronchiectasis | 1(1%) | 59(40%) | 13(57%) | 46(37%) | 0.076 |
| 6.Consolidation | 112(77%) | 29(20%) | 9(39%) | 20(16%) | **0.010** |
| 7.GGO | 0 | 8(5%) | 1(4%) | 7(6%) | 1.000 |
| 8.Mosaic attenuation | 0 | 14(9%) | 1(4%) | 13(10%) | 0.697 |
| Anemia | 102(65%) | 43(40%) | 9(53%) | 34(37%) | 0.228 |
| Elevated CRP | 137(88%) | 23(26%) | 6(50%) | 17(22%) | **0.043** |

Among the 18 patients diagnosed with CPA at baseline, one was lost to follow-up during treatment, and 8 (44%) were reclassified as non-CPA at the end of therapy without antifungal treatment. At the third month of anti-tubercular treatment, five patients had persistent symptoms, with radiological resolution in two (5 out of 8 with available lung imaging) but all having anti-*Aspergillus* IgG levels. At the end-of-TB therapy, six showed symptom resolution, one had cavity resolution, and three had normal radiology. Remarkably, five of the 8 patients (63%) seroconverted, being labelled as ‘self-resolving CPA’. The remaining nine patients (50%) had ‘persistent’ CPA. Additionally, 14 patients developed CPA over time (‘newly diagnosed CPA’). One hundred twenty-seven patients (80%) were never diagnosed with CPA and labelled as ‘never CPA.’

The changes in titre of anti-*Aspergillus* IgG between the three groups (arbitrarily divided into 0-13.59, 13.6-27, >27) are shown in figure 2b.

Among 14 ‘new-CPA’ patients, three patients had symptoms at baseline, only one patient had positive *anti-Aspergillus* IgG serology. At the third month assessment, out of these 14 patients, five had seroconversion (from negative to positive serology). At the end-of-therapy, the remaining nine also seroconverted. Radiologically, four of these patients had cavities at baseline, increasing to seven by the end of therapy. Pleural thickening, a new finding, developed in three patients (out of 14), was observed at the end-of-therapy. The change in cliniico-radio-serological parameters in the groups whose label changed (from CPA to non-CPA or in reverse) is depicted in figures 2c and 2d.

It was found, on logistic regression, that baseline anti-*Aspergillus* IgG titre (OR 1.1, p = 0.002, 95% CI 1.04-1.18) and SGRQ score at third month (OR 1.06, p=0.031, 95% CI 1.005-1.127) predicted the occurrence of CPA at the end-of-therapy.

## Discussion

This study, to the best of our knowledge, is the largest prospective study, examining serially the incidence of CPA and the changes in the anti-*Aspergillus* IgG titre during the course of anti-tubercular therapy in newly diagnosed P.TB patients. This study showed that a significant proportion of P.TB patients (both sputum positive and negative) have concomitant IgG *Aspergillus* positivity at baseline (some fulfilling criteria for CPA) and that the percentage increased progressively during the course of therapy. Interestingly, IgG *Aspergillus* titre (positivity or negativity) and the label of CPA changed in a number of patients from baseline through the course of anti-tubercular therapy.

This study adds to the existing data on co-occurrence of pulmonary aspergillosis with active pulmonary tuberculosis – both clinico-radiologically and microbiologically diagnosed.^8, 15, 16, 17, 18, 19, 9^. Table 3 describes the available literature on the frequency of CPA at various time points in active P.TB patients.

**Table 3:** Different literatures on co-occurrence of CPA at various time points since P.TB diagnosis.

| S No | Author's Name | Country/ Year | Mode of diagnosis of PTB | Study type | Time-point | Estimate of CPA | others |
| --- | --- | --- | --- | --- | --- | --- | --- |
| 1 | Setianingrum et al. <sup>25</sup> | Indonesia/ 2021 | 91/216 (42%) microbiological (TB PCR & or AFB) 91/216 (42%) Clinical, even after performing diagnostics, 34/216 (16%) clinical without performing diagnostics | Prospective, Longitudinal cohort study | Baseline and end-of-therapy | Baseline: proven- 6% probable- 8% possible- 7%<br>End-of-therapy: proven- 8% probable- 5% possible- 3% | Lost to follow-up: 88/216=41 % |
| 2 | Changwhan Kim et al. <sup>26</sup> | Korea/ 2022 | 345/345 (100%) culture-positive pulmonary TB | Retrospective analysis of protocol-based collective data | Baseline and end-of-therapy for serological grouping and then follow up for a median of 25.8 months | Median of 13.5 months after treatment completion: Total- 10/345 (2.9%) Seroconversion group- 7/24 (29.2%) Persistently seropositive group- 3/73 (4.1%) | Significant association with seroconversion of anti-*Aspergillus* IgG and diabetes mellitus with CPA development |
| 3 | Fei Wang et al. <sup>27</sup> | Wuhan pulmonary hospital, China / 2022 | Positive results from sputum or bronchoalveolar lavage fluid mycobacterial culture or nucleic acid amplification test | Retrospective case-control study | Single time point, at the time of recruitment. | Suspected and clinical diagnosis of CPA= 216/9365 (2.3%) PTB+ suspected CPA= 96/216 (44%) PTB+ clinical diagnosis of CPA = 120/216 (56%) Case group, EPTB+CPA= 58/216 (27%) | Pulmonary cavity, COPD and bronchiectasis are significantly associated with development of CPA in etiology-positive PTB (EPTB) group |
| 4 | Abdurakhim Toychiev et al. <sup>28</sup> | Uzbekistan / 2022 | 140/200(70%) smear positive 60/200(30%) smear negative, clinically diagnosed | Prospective observational study | Single point of time- At baseline | CCPA- 13/200 (6.5%) Subacute invasive aspergillosis- 8/200 (4%) Chronic fibrosing pulmonary aspergillosis – 4(2%) | Chronic pulmonary aspergillosis was detected in 11.4% and 20.0% of smear-positive and smear- |
|  |  |  |  |  |  | Single aspergilloma – 3(1.5%) | negative group |
| 5 | Nousheen Iqbal et al.<sup>29</sup> | Pakistan/ 2016 | NA | Retrospective study | Single time point – at recruitment | CPA - 47/69(68.1%)<br>CCPA- 21/47 (44.7%)<br>Simple aspergilloma- 26/47 (55.3%) | DM, mean LOS and hypoxic respiratory failure were identified as independent risk factors of mortality |
| 6 | Waqas Akram et al.<sup>30</sup> | Pakistan/ 2021 | NA | Retrospective study | Single time point – at recruitment | Simple aspergilloma- 122/218(56%)<br>CCPA- 68/218 (31.2%)<br>SAIA-28/218 (12.8%) | PTB was found to be the most common associated lung disease (with n = 137, 62.8%).<br>96/137(70%) -Past TB<br>41/137 (29.9%) - Active TB |

However, barring a single study,^8^ no other study had reported sequentially the incidence of CPA and IgG *Aspergillus* positivity in newly-diagnosed patients with P.TB. Unlike the previous study,^8^ our study reported only proven CPA (but not possible or probable CPA) and still found the proportions comparable (7% at baseline and 14.6% at end-of-therapy of proven CPA) to that reported by Setianingrum et al (7.9% at baseline and 13.3% at end-of-therapy of combined proven and probable CPA).^8^

However, the significance of diagnosis of such CPA in P.TB patients is yet to be ascertained. This is borne by the presence of ‘reversible’ or ‘self-resolving’ CPA – around 44.4 % (95% CI: 21.5% to 69.2%) of newly-diagnosed P.TB patients (6/11 in sputum positive and 2/7 in sputum negative), where the label of CPA was reversed by the end-of-TB-therapy. Since a significant proportion of these patients had resolution of lung lesions on anti-tubercular therapy, significant decrease in anti-*Aspergillus* IgG titre and clinical improvement without any antifungals – it is possible that the anti-tubercular therapy itself led to an indirect effect on *Aspergillus* lung infection. Whether this is due to the partial reduction in lung lesions (e.g. decrease in cavity size) leading to decreased ‘space’ for *Aspergillus* to continue to sustain itself or by hitherto undescribed ‘cross-talk’ between the microbes, needs further research. The APICAL study ^8^ had reported 7/10 cases of CPA to ‘self-resolve’ by the end-of-therapy while Kim et al^9^ had found ‘seroreversion’ (IgG *Aspergillus* converting from positive to negative by end-of-therapy) in 16/345 or 4.6% of patients. In the latter study, there was higher resolution rate of primary cavity after TB treatment in the ‘seroconversion’ group (40%) and ‘persistently seronegative’ group (75.9%) as compared to ‘seroreversion’ group (0%) and ‘persistently seropositive’ group (22.7%).^9^ In our study 5/8 patients (~63%. 95% CI: 24.5-94.5) who had ‘self-resolving CPA” had resolution of their consolidation (without development of new cavity) at the end-of-therapy as compared to their baseline chest images.

In our cohort hemoptysis was commoner in the CPA group at baseline (19.4% vs 13.2%) and at end-of-therapy (9% vs 1%) while cough was commoner at end-of-therapy (43% vs 16%) but not at the baseline. Though SGRQ was comparable in the CPA and non-CPA group at baseline, it was found to decrease significantly in non-CPA group as compared to CPA at end-of-therapy (34.4 vs 53.4, p= 0.0017). This underlines the importance of thorough symptom screening in P.TB patients particularly at follow-up. Chronic and persistent symptoms on follow-up can indicate the presence of CPA in these patients.^20, 21^ CPA is one of the most important post-tuberculous lung diseases which together or in isolation can lead to poor QOL.^22, 23^ Our study suggests, P.TB patients with persistent poor QOL need to be evaluated for CPA, among other complications.

Our study had a few notable limitations. Around 38% of patients could not be followed up at the end-of-therapy despite repeated telephonic reminders, chiefly attributed to financial constraints. Though significant, the percentage (of loss to follow-up) was comparable to that reported previously.^8^ Secondly, limited availability of baseline CT thorax images for some patients may have led to misidentification of lung lesions and potential mislabeling of CPA cases. Thirdly, a small percentage of our patients had their sputum examined for fungal elements because of non-productive cough. Since these patients were not subjected to bronchoalveolar lavage, it is possible a fraction of CPA patients, who were IgG *Aspergillus* negative (~18% of CPA patients have been previously reported from India to have IgG *Aspergillus* negativity)^24^, have been ‘missed’. However, we believe that this (along with aspergilloma on CT with negative IgG *Aspergillus* in 3 patients who were labeled as non-CPA) could have led to underestimation of the proportion of our study patients with CPA. Lastly, since culture confirmation of mycobacterial infection was present in only a small percentage of sputum-negative P.TB patients in our study, it is difficult to infer whether these patients had only CPA or concomitant CPA with P.TB. While 2 out of 7 (28.6%) sputum-negative P.TB patients improved only on anti-tubercular therapy (likely representing true cases of combined P.TB and CPA), 5 patients (71.4%) were cases of ‘persistent CPA’ (likely representing ‘misdiagnosis’ cases).

## Conclusions

We infer from our study that CPA might coexist with or complicate newly diagnosed P.TB patients from the time of diagnosis till the end-of-therapy. Efforts should be made to identify and evaluate patients with persistent symptoms of P.TB patients on therapy, for CPA. Further studies are needed to identify those cases of CPA who might need antifungal treatment as well as those who are likely to self-resolve.

## Data Availability

All data produced in the present study are available upon reasonable request to the authors

## Acknowledgements

None

## Contribution

Author 1: Dhouli Jha

- Collected the data
- Wrote the paper

Author 2: Umesh Kumar

- Contributed data

Author 3: Ved Prakash Meena

- Contributed data or analysis tools

Author 4: Prayas Sethi

- Contributed data or analysis tools

Author 5: Amandeep Singh

- Contributed data or analysis tools

Author 6: Neeraj Nischal

- Contributed data or analysis tools

Author 7: Pankaj Jorwal

- Contributed data or analysis tools

Author 8: Surabhi Vyas

- Contributed data or analysis tools

Author 9: Gagandeep Singh

- Contributed data or analysis tools

Author 10: Immaculata Xess

- Contributed data or analysis tools

Author 11: Urvashi B Singh

- Contributed data or analysis tools

Author 12: Sanjeev Sinha

- Contributed data or analysis tools

Author 13: Anant Mohan

- Contributed data or analysis tools

Author 14: Naveet Wig

- Contributed data or analysis tools

Author 15: Sushil Kumar Kabra

- Contributed data or analysis tools

Author 16: Animesh Ray

- Conceived and designed the analysis
- Contributed data or analysis tools
- Performed the analysis
- Contributed in writing the paper

